# Harnessing Pathology Foundation Models to Accelerate Lymphoma Diagnosis Through Automated Immunohistochemistry Triage

**DOI:** 10.64898/2026.08.11.26360085

**Authors:** Ibrahim Safa, Michael Hazoglu, Pallavi Galera, Chad Vanderbilt, Ali Kamali, Gregory Goldgof, Harini Veeraraghavan, Jue Jiang, Orly Ardon, Luke Geneslaw, Anyi Li, Meng-Lei Zhu, Ahmet Dogan

## Abstract

Pathologic diagnoses of hematopoietic diseases require immunohistochemistry (IHC) stains selected by pathologists upon preview of H&E-stained slides. This multi-step workflow can delay diagnostic turnaround time by days. Hence, we developed the Hematopathology Automatic Triaging System (HATS), which automates IHC panel ordering directly from H&E whole-slide images using pretrained pathology foundation model representations combined with attention-based multiple-instance learning. After the most comprehensive evaluation of pathology foundation models for hematologic malignancy classification to date, encompassing seven publicly available models, we trained HATS on 4,996 whole-slide images from 1,607 patients spanning the ten most common lymphoma diagnostic categories. HATS achieves 84% case-level subtype classification accuracy (0.962 ROC-AUC), translating to 92% IHC panel ordering accuracy. In a blinded reader study, HATS outperforms practicing pathologists at predicting lymphoma subtypes from morphology alone (85% vs 65%). In an independent real-world validation of 230 clinical cases, after directing 7 cases with scant tissue for manual review, HATS-ordered IHC panels were sufficient for diagnosis in 72.6% of cases. By automating the triaging step while preserving full pathologist oversight, HATS offers a safe and practical entry point for clinical AI adoption in pathology.

## INTRODUCTION

Recent innovations in deep learning and artificial intelligence (AI) technologies have resulted in rapid growth of AI applications in healthcare [1]. Digital pathology is well suited for AI-driven clinical applications, particularly in cancer diagnosis and tissue biomarker analytics [2]. Many studies have shown that AI models reproduce histopathologic diagnoses routinely performed by pathologists in many aspects of pathology workflows, such as tumor diagnosis, subtyping, and grading in multiple tumor types [3–10]. Recently, several studies have applied deep learning to lymphoma diagnosis from H&E-stained tissue, ranging from binary tasks such as distinguishing follicular lymphoma from reactive hyperplasia [11] to multiclass subtype classification across three to six diagnostic categories [12–15]. Self-supervised pretraining on pathology images has also been explored for molecular prediction in large B-cell lymphoma [16], though these efforts predate the emergence of large-scale pathology foundation models. More broadly, these approaches have largely tackled isolated diagnostic tasks rather than the full diagnostic workup that hematopathology requires.

The diagnostic workflow in hematopathology is particularly well suited for AI-assisted triage, as reaching a precise diagnosis requires not only morphologic review but also ancillary tests such as immunohistochemistry (IHC), flow cytometry, cytogenetic, and molecular studies. The typical workflow for hematopathology includes the following sequential steps (Fig. 1a): Hematoxylin and eosin (HE) slides preparation, pathologist preview of slides and ancillary tests, IHC ordering, final pathologist review, diagnosis and reporting. In some complex cases, the pathologist cannot make a final diagnosis based on the HE and IHC stains ordered in the first round; then, additional IHC stains are ordered. Pathologist previews and IHC orders typically require an additional turnaround day. Overall, routine hematopathology workflows require triaging in more than 90% of specimens to order appropriate IHC panels for further differential diagnosis. This process is not only time-consuming for pathologists but also demands significant administrative and laboratory coordination to order and distribute slides, further delaying final diagnoses and increasing costs. Prompt and accurate triage of IHC orders is therefore key to reducing turnaround time and increasing throughput. While most clinical AI applications require near-perfect accuracy because they directly inform diagnostic or therapeutic decisions, triaging is inherently more tolerant of error: an unnecessary IHC stain will be caught during pathologist’s review before any clinical decision is made. This makes triaging uniquely suited for early AI adoption, enhancing workflow efficiency without compromising diagnostic safety. Large-scale foundation models have transformed computational pathology by learning visual patterns from massive datasets that generalize across diagnostic tasks[17]. These models are pretrained on millions of unlabeled histopathology images using self-supervised learning (SSL), which enables the learning of visual and spatial tissue structure as reusable numerical representations without requiring expert labels. In this work, we conducted the most comprehensive evaluation of pathology foundation models for hematologic malignancy classification to date, benchmarking seven major publicly available models which differ in their architectures, pretraining strategies, and training data sources: Prov-GigaPath, a vision transformer trained on 1.3 billion image tiles from over 170,000 whole-slide images[18]; CONCH, a vision-language model jointly trained on 1.17 million image-caption pairs[19]; MUSK, which aligns 50 million pathology images with one billion text tokens for precision oncology[20]; Virchow2, a 632-million parameter network trained on 3.1 million WSIs spanning over 40 tissue types[21]; H-optimus-0, a 1.1-billion parameter vision transformer trained on over 500,000 WSIs across multiple tissue types[22]; UNI v2, a vision transformer trained on over 100,000 histopathology slides from diverse sources [23]; and Phikon v2, an open source DINOv2-based model pretrained on TCGA and other public histopathology datasets. Each model produces patch-level embeddings ranging from 512 to 1,536 dimensions [24].

**Figure 1.**
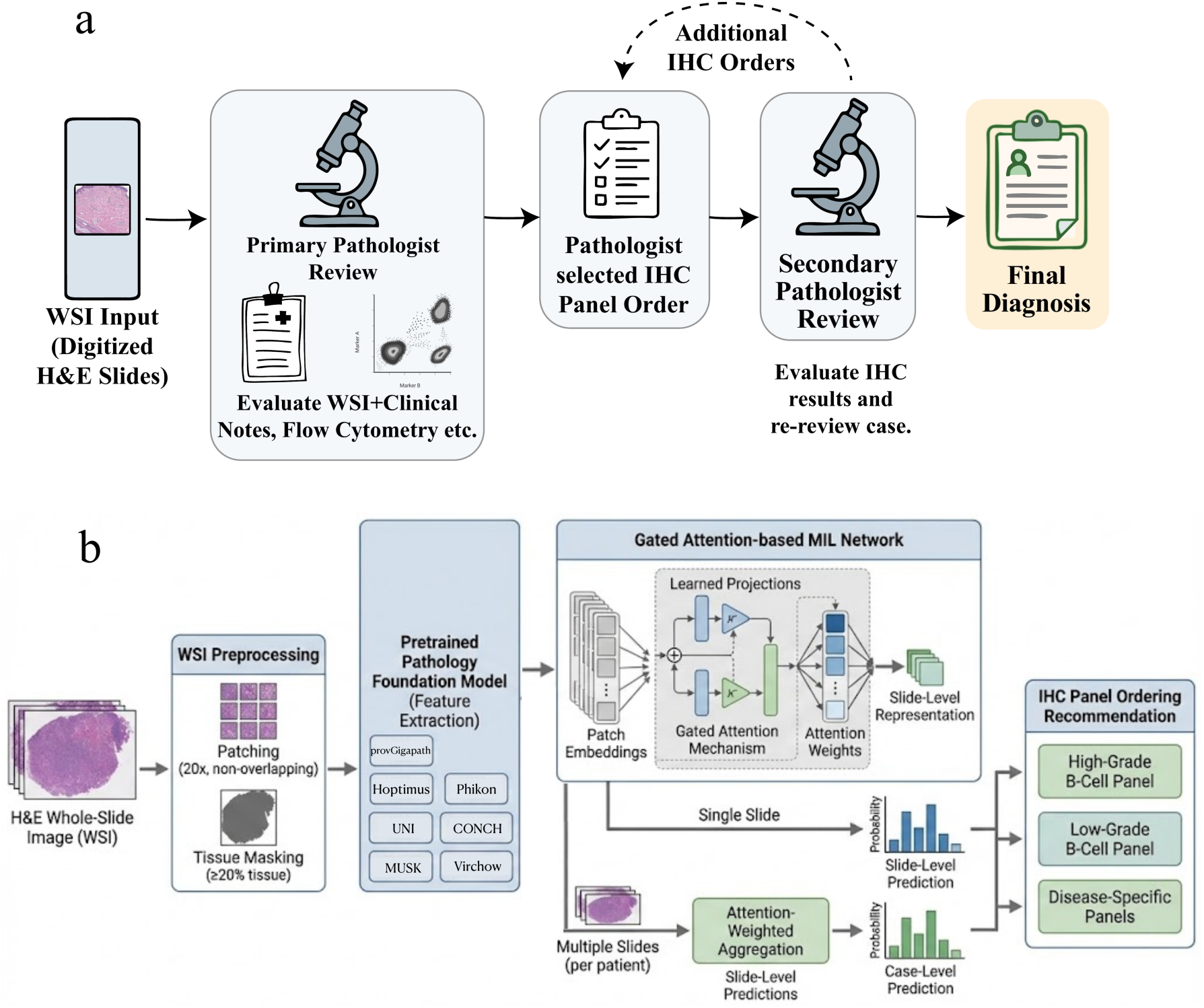
**(a) Current Hematopathology diagnostic workflow.** Schematic showing the current workflow where a pathologist is required to review a case multiple times over several days in order to reach a final diagnosis. An initial review is done to study the WSI, along with clinical notes and other ancillary data. The pathologist then has to review the case at least once more to reach a diagnosis. In some cases, further IHC tests need to be ordered after the second round. **(b) HATS pipeline overview.** Schematic illustrating the end-to-end HATS workflow from whole-slide image to IHC panel ordering. H&E-stained whole-slide images are first preprocessed by dividing into non-overlapping patches at 20× magnification with tissue masking to exclude background regions. A pretrained pathology foundation model (H-optimus-0) extracts feature embeddings from each patch. These embeddings are processed by a gated attention-based multiple-instance learning (MIL) network, which outputs a probability distribution across ten lymphoma subtypes. When multiple slides are available for a patient, slide-level predictions are aggregated into a single case-level classification using attention-weighted averaging. Finally, the predicted subtype is mapped to the appropriate IHC panel (high-grade B-cell, low-grade B-cell, or disease-specific panels), which is automatically ordered for pathologist review.

We combine these patch-level representations with an attention-based deep learning approach [25], in which the model learns to identify and prioritize the most informative regions of a slide to produce a single diagnostic prediction. Using this approach, we developed the Hematopathology Automatic Triaging System (HATS), a framework that predicts IHC panel orders directly from H&E whole-slide images. Here, we show that HATS achieves high diagnostic accuracy across ten hematopathologic categories, with strong performance translating to over 90% IHC panel ordering accuracy on a held-out test set. In a real-world clinical evaluation, HATS demonstrates the ability to reduce diagnostic turnaround time while preserving pathologist oversight.

## RESULTS

### Model design

Because hematopoietic malignancies infiltrate diffusely throughout the tissue rather than forming localized masses, large parts of the H&E slide are potentially relevant for diagnosis. HATS was therefore designed around two complementary strengths: pretrained foundation models, which provide a robust backbone encoding generalizable histological patterns, and our institution’s large hematopathology case archive, which supplies the disease-specific training volume needed to refine these representations into a clinically effective classifier.

The HATS pipeline proceeds in three stages (Fig. 1b). First, each whole-slide image is divided into non-overlapping patches, and a frozen foundation model encodes each patch into a high-dimensional feature vector capturing numerical representations of visual features. Second, these patch-level features are aggregated into a single slide-level representation using attention-based multiple instance learning (AbMIL). In this framework, each slide is treated as a “bag” of patch instances, and a gated attention mechanism learns to assign diagnostic relevance scores to each patch, enabling the model to focus on morphologically informative regions while down-weighting background or uninformative tissue. Because foundation model weights remain frozen, only the attention-based classifier undergoes training. Third, for patients with multiple slides from the same case, slide-level predictions are combined to produce a final case-level classification across the ten lymphoma subtypes. Notably, we systematically compared major publicly available pathology foundation models (UNI v2, Prov-GigaPath, CONCH, MUSK, Virchow2, H-optimus-0, and Phikon v2) using identical downstream classifiers to identify the optimal feature representation for this task. Detailed results of this comparison are presented in a separate section below. H-optimus-0 yielded the strongest performance and was selected for all subsequent analyses.

The study cohort comprised 4,996 whole slide images from 1,607 patients spanning ten diagnostic categories: follicular lymphoma (FL), diffuse large B-cell lymphoma (DLBCL), plasma cell neoplasm (PCN), marginal zone lymphoma (MZL), mantle cell lymphoma (MCL), classical Hodgkin lymphoma (CHL), chronic lymphocytic leukemia/small lymphocytic lymphoma (CLL), anaplastic large cell lymphoma (ALCL), angioimmunoblastic T-cell lymphoma (AITL), and Burkitt lymphoma (BL). The class distribution reflects real-world clinical prevalence in a tertiary cancer center. The dataset was partitioned at the patient level to prevent data leakage. Using this cohort, we evaluated HATS classification performance at both the slide and case levels.

### HATS performance

Our H-optimus-0-based model demonstrates robust slide-level diagnostic performance across ten histopathologic categories. It achieved a macro-averaged ROC AUC of 0.962 ± 0.009 and area under the precision-recall curve (AUPRC) of 0.792 ± 0.034 on the held-out test set. At the slide level, the model achieved accuracy of 79.4% ± 1.5%, precision of 76.0% ± 3.5%, recall of 69.6% ± 5.3%, and F1-score of 70.8% ± 3.7% (macro-averaged). All values represent mean ± 1 standard deviation on held out cross-validation test sets (Fig. 2a-c).

**Figure 2.**
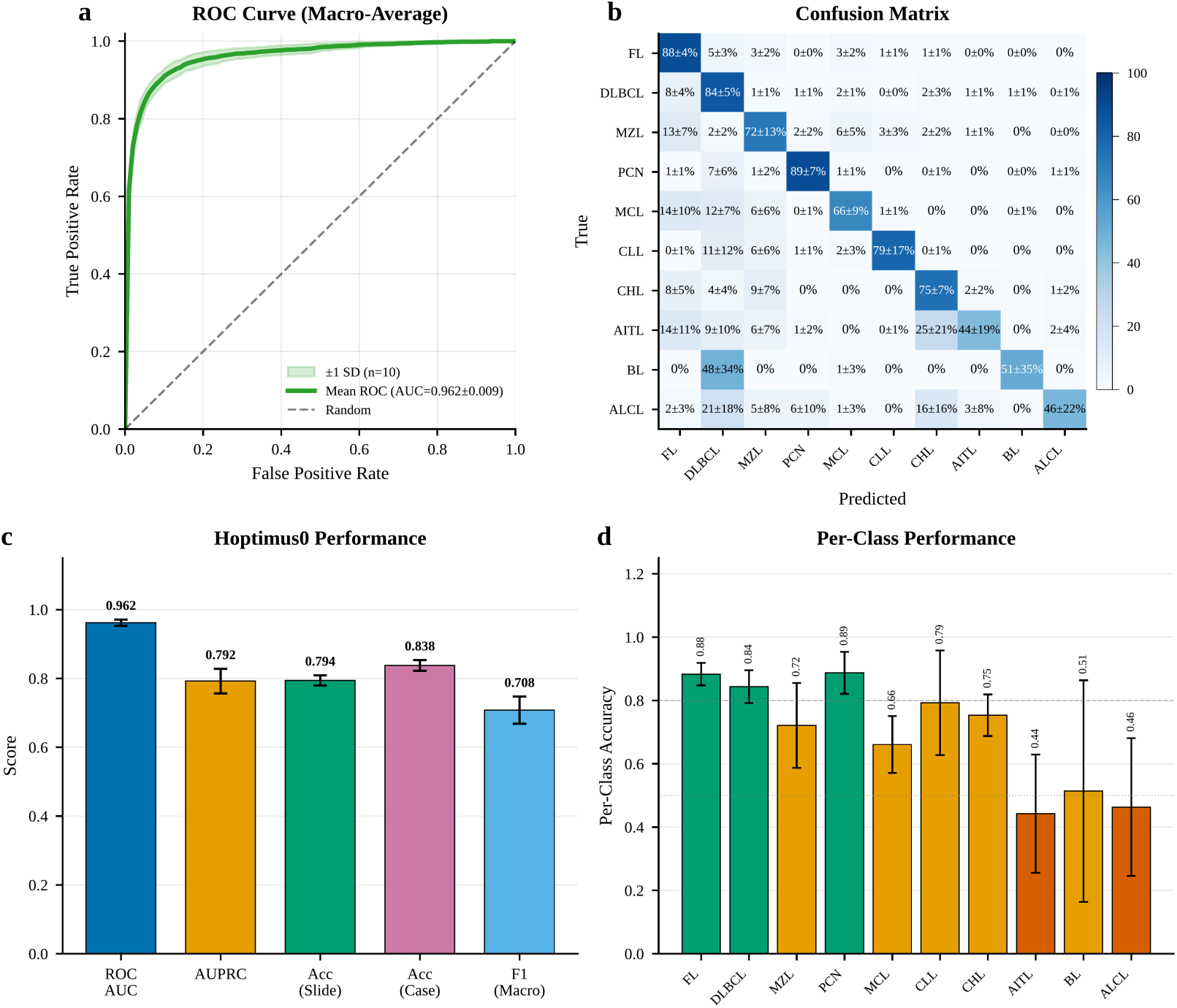
HATS classification performance using H-optimus-0 features. **(a)** Receiver operating characteristic (ROC) curve showing macro-averaged true positive rate versus false positive rate. Shaded region indicates ±1 standard deviation across 10 independent training runs with different data splits. Dashed line indicates random classifier performance. **(b)** Normalized confusion matrix showing classification accuracy for each lymphoma subtype. Values represent mean percentage ± standard deviation across seeds. Classes are ordered by prevalence in the test set (FL most common, ALCL least common). **(c)** Summary of test set performance metrics across all seeds. Error bars indicate standard deviation. ROC AUC, area under the receiver operating characteristic curve; AUPRC, area under the precision-recall curve; Acc (Slide), slide-level accuracy; Acc (Case), case-level accuracy using attention-weighted aggregation; F1 (Macro), macro-averaged F1 score. **(d)** Per-class classification accuracy (equivalent to recall). Bar colors indicate performance level: green (≥80%), orange (50–80%), red (<50%). Error bars indicate standard deviation across seeds.

Clinical cases typically include multiple slides from the same biopsy, all carrying the same diagnosis. To leverage this redundancy, we aggregated slide-level predictions into case-level classifications using four strategies: entropy-weighted aggregation, max-pooling, mean-pooling, and majority voting (Supplementary Fig. 1). All four improved over slide-level accuracy (79.4% ± 1.5%), achieving case-level accuracies of 83.8% ± 1.5%, 83.6% ± 1.9%, 83.8% ± 1.4%, and 83.5% ± 1.3%, respectively. The narrow spread across methods suggests that slide-level predictions within a case were generally consistent. We adopted entropy-weighted aggregation for all subsequent analyses because, in hematologic malignancies where disease involvement is typically uniform across tissue sections, slides producing relatively focused attention patterns are more likely to contain clear diagnostic morphology.

The model outputs a confidence score for each prediction ranging from 0 to 1. This score is well calibrated to prediction quality: only 8% of correct predictions fall below a confidence threshold of 0.6, compared with over 42% of incorrect predictions (Fig. 3d). A threshold-based deferral strategy could therefore reduce erroneous IHC orders by >40% while affecting fewer than one in twelve correct orders. The optimal threshold will depend on institutional error tolerance and workflow constraints.

**Figure 3.**
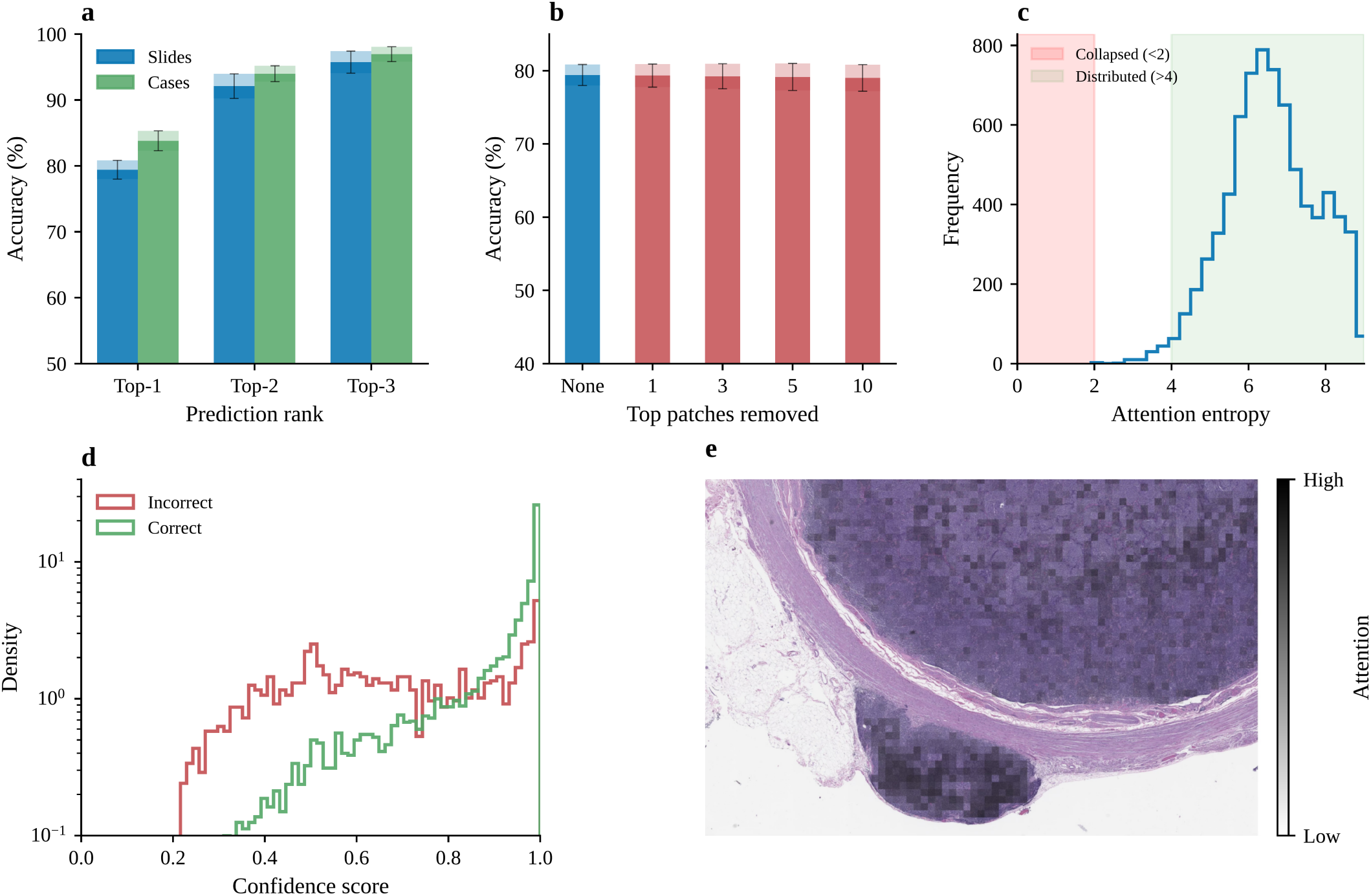
Model robustness and attention analysis. **(a)** Top-k classification accuracy showing how often the correct diagnosis appears among the model’s top predictions. Slide-level (blue) and case-level (green) accuracy are shown for top-1, top-2, and top-3 predictions. Error bars indicate standard deviation across independent training runs. **(b)** Instance ablation test evaluating attention robustness. Bars show classification accuracy when removing the top 1, 3, 5, or 10 highest-attention patches from each slide. Minimal accuracy degradation indicates the model relies on distributed morphological patterns rather than isolated instances. **(c)** Distribution of attention entropy across test slides. Higher entropy indicates attention is distributed across more patches rather than collapsed onto a few instances. Shaded regions indicate interpretive thresholds: collapsed attention (<2) versus well-distributed attention (>4). **(d)** Distribution of confidence scores output by our model comparing correct and incorrect answers, showing a clear distinction between the two. **(e)** Representative section of a whole-slide image with attention overlay. Darker regions indicate higher attention weights, highlighting tissue areas the model considers diagnostically relevant.

Per-class accuracy was better for the more common subtypes (Fig. 2d). The common subtypes demonstrated robust classification: FL (91.2% case-level accuracy), DLBCL (89.8%), and PCN (94.8%) were classified with high accuracy. CHL achieved 80.2% case-level accuracy, and MZL achieved 74.1%. Rarer entities were less reliably classified at the subtype level: AITL, ALCL, and BL each fell below 60%. Though, as described below, many of these cases still received an appropriate IHC panel despite incorrect subtype predictions.

### Model robustness and quality assessment

We performed several analyses to verify that our model learns robust and generalizable morphological features rather than exploiting statistical shortcuts or artifacts. We evaluated top-k prediction accuracy across multiple training runs (Fig. 3a). The true diagnosis appeared within the top two predictions 92.4% ± 1.2% of the time and within the top three 95.9% ± 1.2%, improving to 94.4% ± 0.7% and 97.0% ± 1.2% at the case level. In the clinical workflow, these top-ranked predictions can be presented alongside their confidence scores, effectively narrowing a ten-way differential to a shortlist of two or three candidates for pathologist review.

Another common failure mode in attention-based MIL is “attention collapse” where the model focuses on a small subset of instances, potentially learning to recognize specific artifacts or memorizing signature patches rather than learning distributed morphological patterns [26]. This is especially important in the context of lymphomas, as they are commonly present as diffuse tumor infiltration. Across 10 independent training runs, attention entropy averaged 6.59 ± 0.10, with higher values indicating more distributed attention (Fig. 3c), and the top three highest-weighted patches collectively received only 5.3% ± 0.5% of total attention, with 0% of slides showing any evidence of collapse (Fig. 3c,e). These results confirm that HATS predictions arise from patterns distributed across many tissue regions rather than dependence on isolated instances. Instance ablation testing corroborated this finding. Systematically removing the top one, three, or five highest-attention patches from each slide degraded accuracy by only 0.10%, 0.23%, and 0.33%, respectively (Fig. 3b), confirming that no small subset of patches drives the model’s predictions.

In addition, stratification by tissue type confirmed that HATS achieves consistent performance across lymph node and non-lymph node specimens, supporting the generalizability of the model across the diversity of specimen types encountered in routine practice (Supplementary Note 3; Supplementary Table 4; Supplementary Fig. 4).

### Foundation model comparison

We conducted the most comprehensive evaluation of pathology foundation models for hematologic malignancy classification to date. All models were tested using identical training procedures with a gated attention-based MIL classifier and multi-seed experiments to account for variability in initialization and data partitioning. H-optimus-0 achieved the best overall performance with a mean slide-level accuracy (on the held-out test set) of 79.4% ± 1.5%, case-level accuracy of 83.8% ± 1.5%, and ROC AUC of 0.962 ± 0.009 (all ± SD). UNI v2 achieved slide-level accuracy of 77.0% ± 2.6%, case-level accuracy of 79.4% ± 2.3%, and ROC AUC of 0.955 ± 0.012. Virchow2 achieved slide-level accuracy of 76.7% ± 2.8%, case-level accuracy of 80.0% ± 2.9%, and ROC AUC of 0.955 ± 0.010. Prov-GigaPath achieved slide-level accuracy of 75.3% ± 2.8%, case-level accuracy of 78.3% ± 2.9%, and ROC AUC of 0.949 ± 0.015. CONCH achieved slide-level accuracy of 71.0% ± 3.1%, case-level accuracy of 73.9% ± 3.3%, and ROC AUC of 0.942 ± 0.009. MUSK achieved slide-level accuracy of 69.3% ± 2.2%, case-level accuracy of 72.7% ± 3.4%, and ROC AUC of 0.929 ± 0.009. Phikon v2 demonstrated the lowest performance with slide-level accuracy of 68.2% ± 2.5%, case-level accuracy of 71.1% ± 2.5%, and ROC AUC of 0.928 ± 0.011(Fig. 4a-c). More details are included in the Supplementary Materials (Supplementary Note 1, Supplementary Tables 1-2; Supplementary Fig. 2).

**Figure 4.**
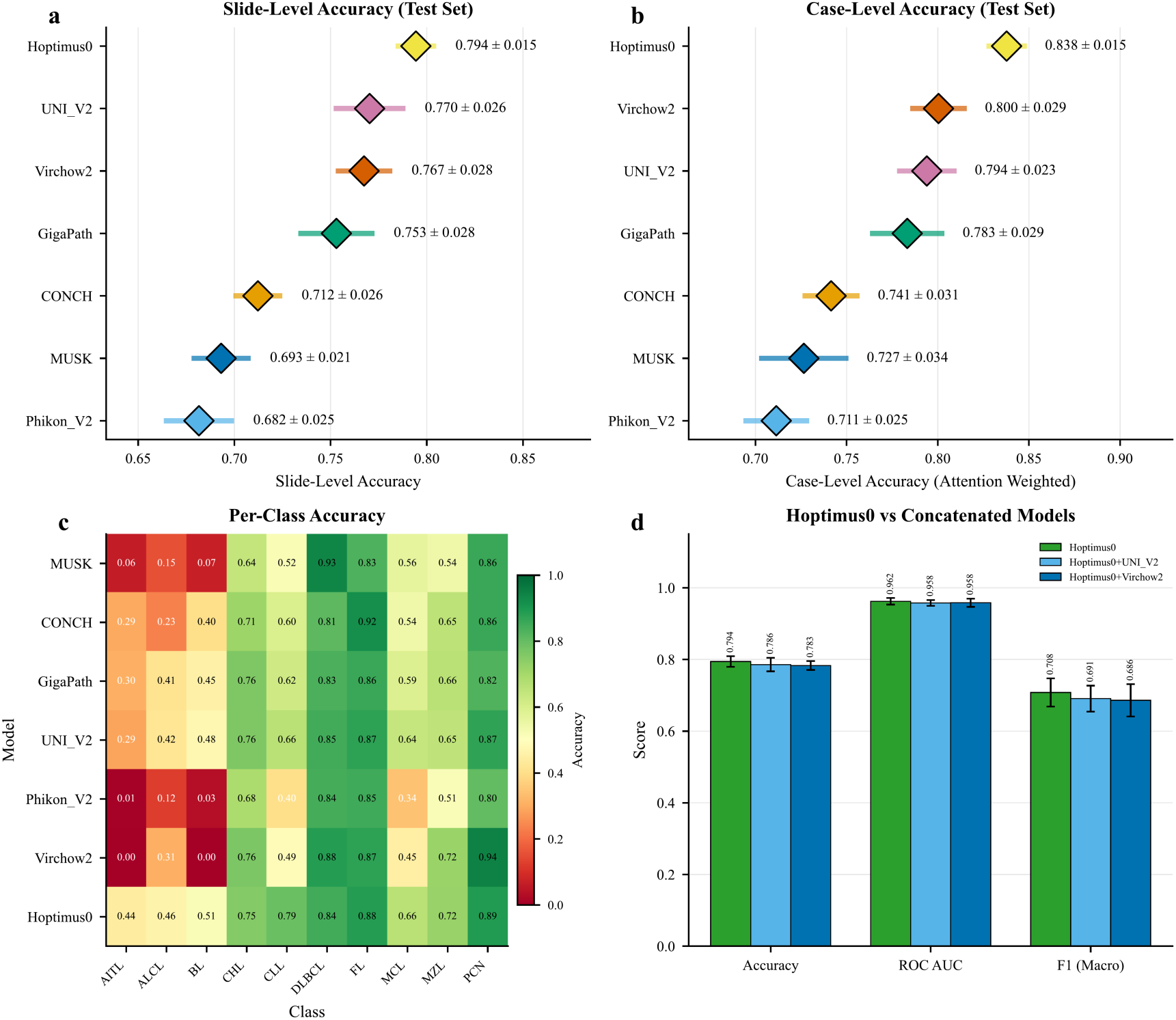
Comparison of foundation models for lymphoma classification. **(a)** Forest plot showing slide-level test set accuracy for each foundation model. Diamonds indicate mean accuracy; horizontal lines indicate 95% confidence intervals. Values shown as mean ± standard deviation. **(b)** Forest plot showing case-level test accuracy using attention-weighted aggregation. **(c)** Heatmap showing per-class accuracy for each foundation model. Color scale ranges from 0 (red) to 1 (green). **(d)** Comparison of H-optimus-0 alone versus ensemble models combining features from multiple foundation models. Neither ensemble approach outperformed H-optimus-0 alone, suggesting overlapping rather than complementary feature representations.

We further investigated if a combination of foundation models could improve the final performance. We utilized feature concatenation approaches that combine features from top-performing models. Combining H-optimus-0 and Virchow2 (feature dimension 4096) achieved slide-level accuracy of 77.6%, case-level accuracy of 81.9%, and ROC AUC of 0.954. This concatenation approach did not improve upon H-optimus-0 alone, suggesting that the evaluated foundation models capture overlapping morphological features rather than complementary information. Based on the superior performance of H-optimus-0 across multiple random seeds, it was selected as the final feature extractor for clinical application (Fig. 4d).

To assess the intrinsic performance of these foundation model embeddings independent of any learned classifier, we performed a parameter-free k-nearest neighbor (k-NN) analysis using H-optimus-0 representations. Each slide’s patch-level embeddings were mean-pooled into a single fixed-dimensional vector, producing a compact slide-level representation without any learned aggregation. Using cosine distance and inverse-distance weighted voting, the k-NN classifier achieved 68% slide-level accuracy on the held-out validation set, well above the 10% expected by chance for a ten-class problem. This result demonstrates that the frozen H-optimus-0 embeddings place same-subtype slides near each other in the representation space even before any task-specific training, confirming that the foundation model captures intrinsic morphological distinctions among lymphoma subtypes. The subsequent addition of the gated attention mechanism through ABMIL training then pushes overall accuracy from 68% to 79% at the slide level. However, our MIL network substantially improves accuracy for rare subtypes compared to the k-NN baseline, suggesting the attention mechanism is particularly important for distinguishing rare subtypes where global mean-pooled representations alone are insufficient (Supplementary Note 2; Supplementary Fig. 3).

### Superior performance by HATS compared with practicing pathologists

To benchmark HATS against pathologists’ performance, we conducted a blinded evaluation in which practicing pathologists reviewed the same H&E slides used in our held-out test set. Pathologists were given access only to H&E slides without clinical notes, flow cytometry, or patient history, all information they would routinely use but which is unavailable to the model. A portal was then built where pathologists could review randomly assigned slides using an embedded WSI digital slide viewer, and submit their classification anonymously (Fig. 5a). We asked participating pathologists to classify these WSIs into one of the 6 major classes using morphology only. The comparison was limited to six major categories, excluding T-cell lymphomas (AITL, ALCL), which are generally not distinguishable by H&E morphology alone, and Burkitt lymphoma and classic Hodgkin lymphoma, for which the small number of cases in the pathologist reviewed set precluded statistically meaningful comparison.

**Figure 5.**
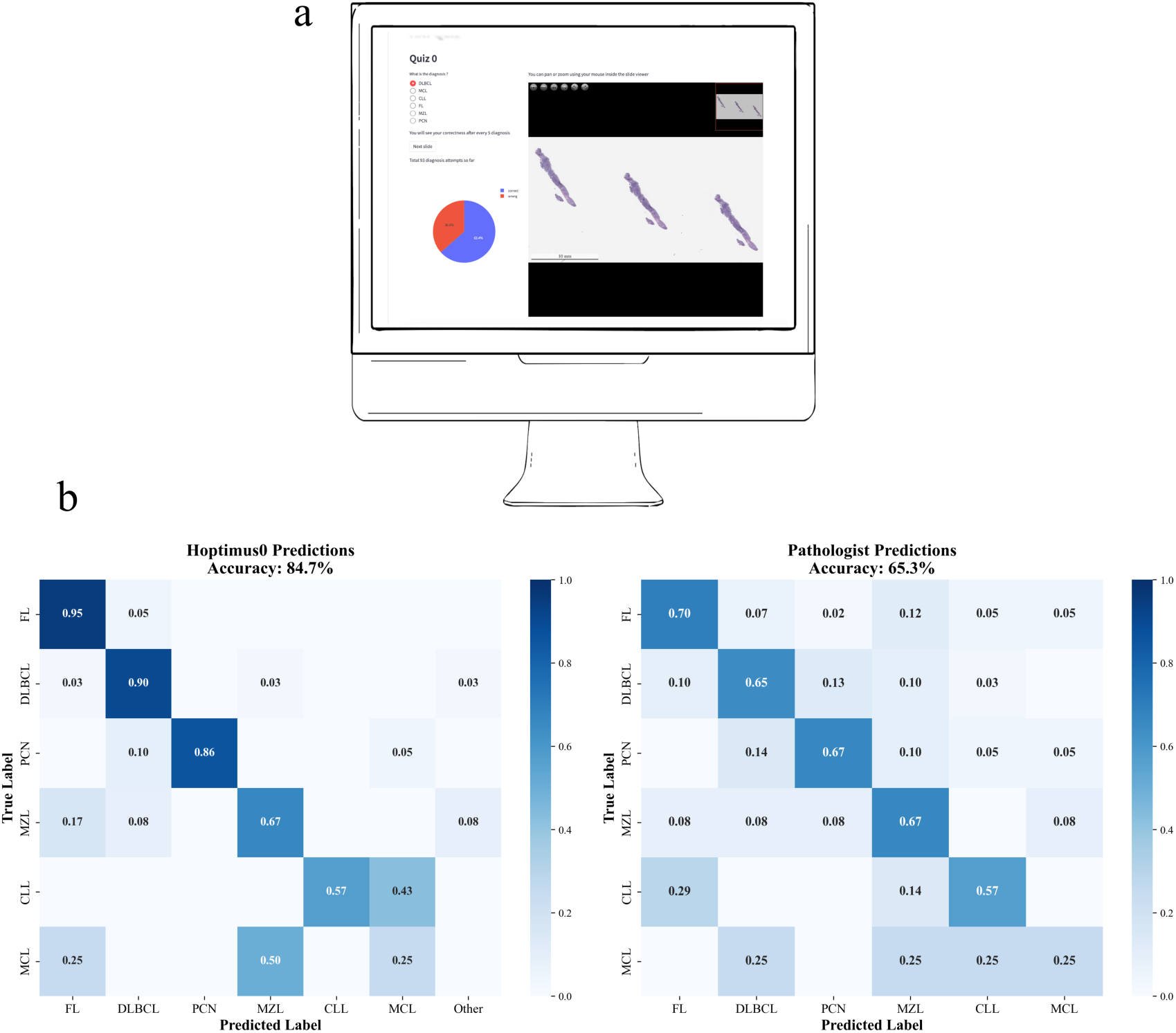
Comparison of AI and pathologist performance on morphology-only classification. **(a)** Screenshot of the web-based review interface used by pathologists to evaluate holdout cases. Pathologists viewed H&E whole-slide images without access to clinical history, flow cytometry, or immunohistochemistry results. **(b)** Left: Confusion matrix showing HATS model predictions on the holdout set (118 slides, 90 cases). Values indicate the proportion of cases with each true label (rows) classified into each predicted category (columns). Overall accuracy: 84.7%. Right: Confusion matrix showing pathologist predictions on the same holdout set. Pathologists achieved 65.3% accuracy when restricted to H&E morphology alone, reflecting the inherent difficulty of lymphoma subtyping without ancillary studies. Classes are ordered by prevalence in the holdout set.

On this matched evaluation, HATS achieved 85% accuracy compared to 65% for pathologists (Fig. 5b,c) after 137 independent pathologist evaluations. This relatively low accuracy reflects the difficulty of lymphoma subtyping from morphology alone, a task where even expert pathologists typically rely on ancillary studies and tests to reach a decision. Rather than suggesting the model replaces pathologist judgment, this result supports the premise of HATS: that AI can perform the morphology-based triaging step at least as reliably as manual preview, freeing pathologists to focus on the integrative diagnostic reasoning.

### AI-assisted workflow and IHC panel ordering

To translate the model’s predictive performance into clinical benefit, we designed an AI-assisted IHC ordering workflow integrated into routine hematopathology practice (Fig. 6). Upon H&E slide scanning, HATS generates probability scores across the ten diagnostic categories and automatically orders one of three IHC panels based on the top prediction: a high-grade B-cell panel for DLBCL, FL, or BL; a low-grade B-cell panel for CLL, MCL, or MZL; or a disease-specific panel for CHL, AITL, ALCL, and PCN (See Supplementary Table 3 for a full list of IHC stains per panel). The high-grade panel differentiates DLBCL, BL, and FL, and enables cell-of-origin determination for DLBCL. The low-grade panel differentiates CLL, MCL, and MZL, and is also sufficient for diagnosing FL. Because the low-grade panel covers FL, an additional rule applies: when the model’s top prediction is FL but its second-ranked prediction is a low-grade category, the low-grade panel is ordered instead, capturing cases where the model is uncertain between FL and a low-grade B-cell lymphoma. This ensures the correct panel is ordered even when the exact subtype prediction is wrong, which substantially improves panel accuracy for the low-grade categories CLL, MCL, and MZL.

**Figure 6.**
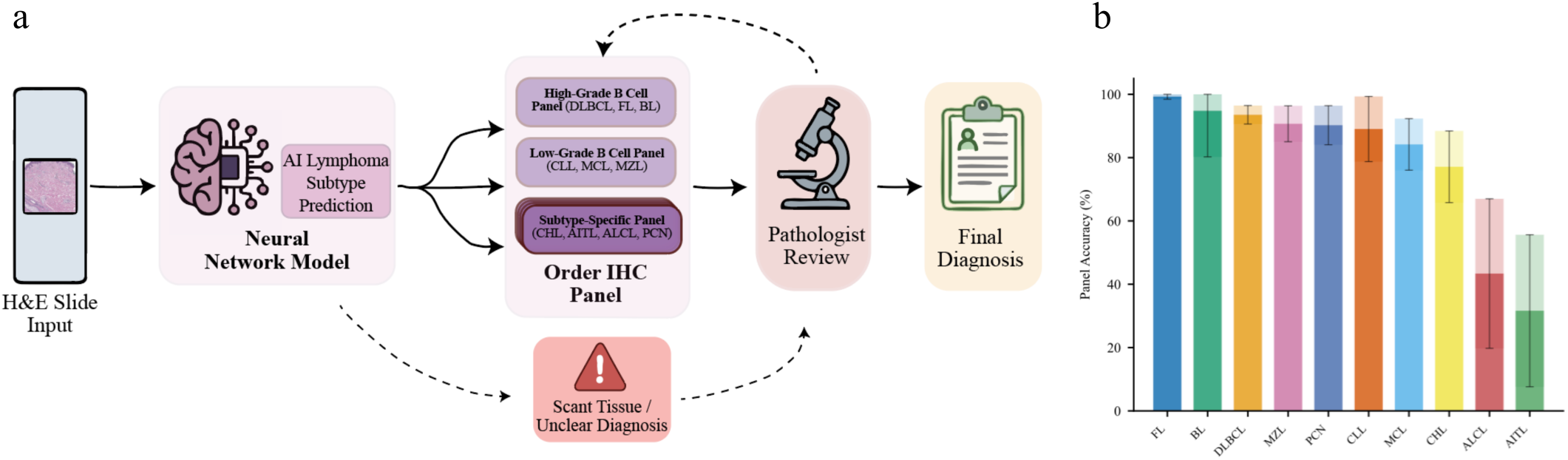
HATS clinical workflow for automated IHC panel ordering. (a) Schematic illustrating the AI-assisted triaging workflow integrated into hematopathology practice. When an H&E whole-slide image is scanned, HATS automatically generates subtype predictions with associated confidence scores before pathologist review. Based on the top prediction, the system orders the appropriate IHC panel: a high-grade B-cell panel (CD20, CD3, CD10, BCL2, BCL6, MUM1, MYC, CD21, CD23, Ki67) for predictions of DLBCL, FL, or BL; a low-grade B-cell panel for CLL, MCL, or MZL; or disease-specific panels for CHL, AITL, ALCL, or PCN. The IHC ordering is automated, eliminating the manual preview step that traditionally adds 1–2 days of turnaround time. The pathologist receives the final package for review: the original H&E slide, the AI prediction with confidence scores and attention metrics, and the resulting IHC stains, enabling efficient and informed diagnostic decision-making while maintaining full human oversight of the final diagnosis. In cases where our model cannot reliably make a prediction (not enough tissue or very low confidence prediction), the case is sent for manual review. (b) Per-class accuracy showing results of the automated panel ordering workflow applied to the test set.

With IHC stains prepared in advance, the pathologist receives the H&E slide, AI prediction report, and completed IHC stains together, enabling a final diagnosis in a single review session rather than the conventional two-step process. If the pre-ordered panel proves insufficient, particularly in challenging or unusual cases, additional stains can always be ordered by pathologists. The final diagnosis remains entirely under pathologists’ control.

We evaluated this workflow by simulating panel ordering on the hold-out test set. The model achieved an overall 92.0% ± 1.8% panel accuracy (Fig. 6b), meaning that in more than 9 out of 10 cases, the automatically ordered IHC panel provides sufficient information for a final diagnosis. Performance was highest for the most common B-cell lymphomas: DLBCL (94.6%), FL (99.5%), MZL (93.2%), and CLL (93.0%). The low-grade B-cell panel demonstrated robust performance for CLL (93.0%), MCL (86.8%), and MZL (93.2%). For individual entity panels, PCN (94.8%) and CHL (80.2%) showed reliable panel assignment. The rare lymphomas AITL (49.8%) and ALCL (50.5%) had lower panel accuracy as expected, reflecting both their scarcity in the training data and the inherent difficulty of identifying T-cell subtypes from morphology alone; these cases would more frequently require pathologist-directed panel selection. A key driver of the higher accuracy is the grouping design: because related subtypes share the same panel, subtype-level misclassifications within a group do not result in incorrect panel orders. For example, a case misclassified between DLBCL and FL still receives the appropriate high-grade B-cell panel.

### Real-world clinical validation

In routine clinical practice, the spectrum of cases encountered extends far beyond the ten categories in our model — encompassing benign conditions, ambiguous or indeterminate diagnoses, rare disease types, cases with multiple concurrent diagnoses, and non-lymphoid neoplasms. To evaluate HATS in a realistic clinical setting, we assembled an independent validation cohort comprising an unfiltered, consecutive series of all tissue specimens directly submitted to the Memorial Sloan Kettering Cancer Center (MSKCC) hematopathology service over a five-month period. The trial set includes 608 WSIs from 223 cases (after excluding 7 cases with scant tissue from total 230 cases). No cases were excluded based on diagnosis, complexity, or specimen quality, ensuring the cohort faithfully represents the full diversity and difficulty of daily clinical practice. Critically, only 59% of the cases fell into one of the ten categories in our model (Fig. 7a). All slides were processed through the identical preprocessing and inference pipeline used during training. Slides with fewer than fifty usable tiles were considered scant and routed to manual pathologist review.

**Figure 7.**
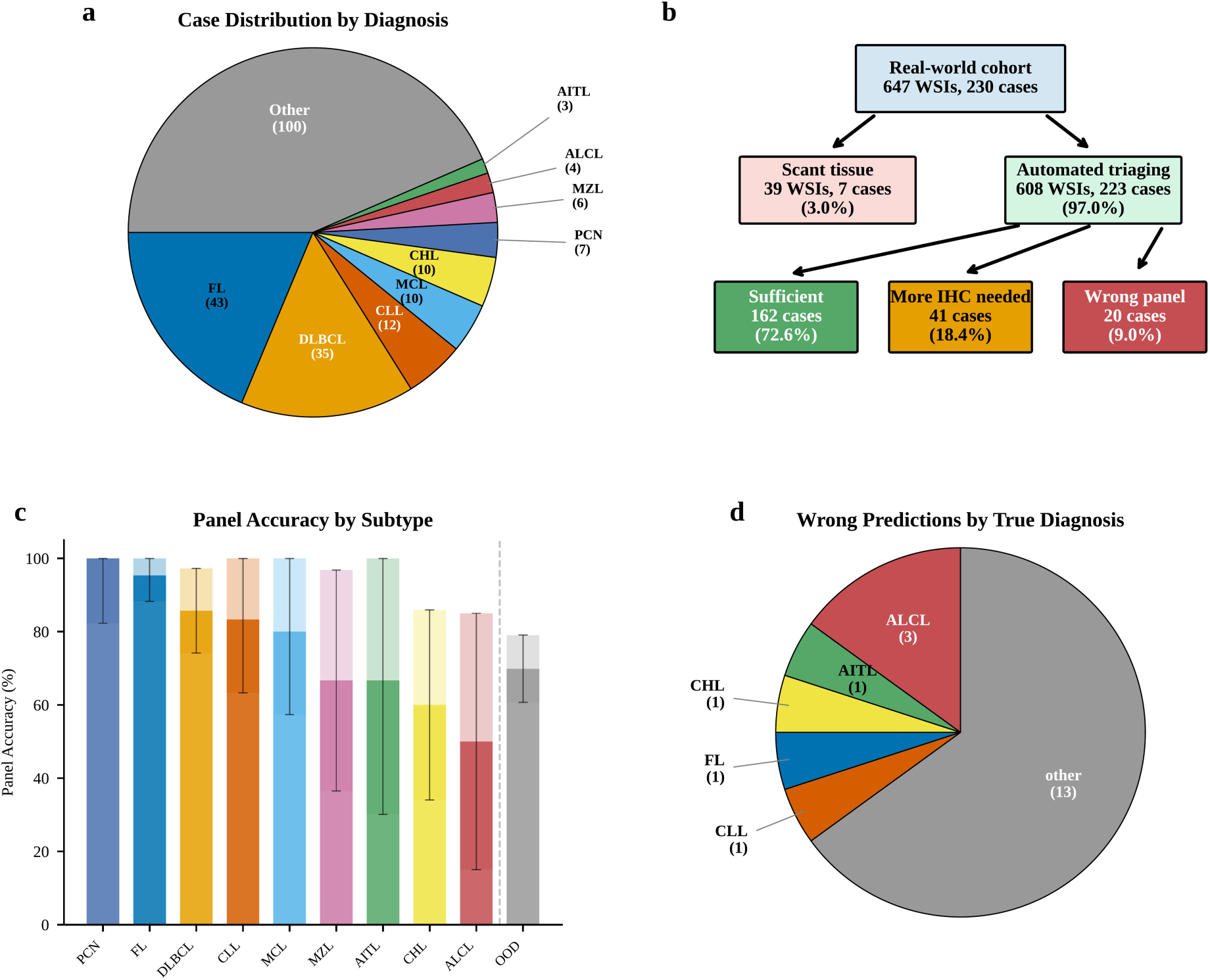
Clinical validation of HATS. Figure displaying the dataset breakdown and performance of our real-world clinical evaluation of HATS. (a) Distribution of clinical trial cases by diagnosis (b) Flowchart summarizing triaging outcomes. Cases with scant tissue were flagged for manual pathologist review; the rest of the cases were classified as either sufficient (green, model order covered the diagnosis), more IHC needed (orange, additional stains required after pathologist review), or wrong panel (red, predicted panels missed the diagnosis). (c) Per-subtype panel accuracy at the case level, ordered by accuracy; bars show mean across 10 random seeds with standard deviation. (d) Distribution of wrong predictions by true diagnosis.

Among slides with a known diagnosis in our ten-category schema, the model achieved an overall panel accuracy of 82% (106/130 cases), consistent with test set performance shown previously. Spearman rank correlation between test-set and real-world per-subtype panel accuracies was strong and statistically significant (rho = 0.72, p = 0.029), indicating that the relative difficulty ordering of subtypes was preserved in the clinical setting.

To assess clinical utility across the full case spectrum, an expert pathologist reviewed the IHC panels assigned by HATS against the final clinical diagnosis for all 223 cases, including those with diagnoses outside of our ten trained subtypes. The study was used to determine when the ordered panels were sufficient to make the final diagnosis upon review, regardless of the predicted subtype. IHC panels ordered by HATS were fully sufficient for diagnosis in 72.6% of the cases; partially sufficient in 18.4% of the cases (requiring additional IHC stains to complete the workup); and incorrect in only 9% (Fig. 7b).

The partially sufficient cases were predominantly driven by complex diagnoses outside our model’s trained categories. Among the incorrectly triaged cases, approximately half involved T-cell lymphomas, which are diagnostically challenging, even for experienced pathologists, in the absence of preliminary flow cytometry data. The remainder included rare entities such as myeloid neoplasms and carcinomas that were outside the scope of the model.

## DISCUSSION

In this study, we developed a clinically deployable workflow that uses AI to support hematopathology triage. Using H&E slides alone, our model achieved high case-level accuracy across ten lymphoma subtypes, demonstrating that morphology contains sufficient signal to guide early decision-making. While a definitive lymphoma diagnosis requires integrating morphology with immunophenotyping and, when indicated, molecular testing, our results suggest that the model exceeds practicing pathologists’ performance on the triaging task based on morphology alone. Importantly, HATS is designed to facilitate, not replace, the diagnostic process: it targets repetitive tasks that consume pathologists’ time, allowing effort to be redirected toward more complex diagnostic activities.

Operationally, the HATS-enabled workflow can generate suggested IHC orders within minutes of slide scanning, enabling near–real time triage. In contrast, traditional manual preview for IHC ordering typically adds additional day of turnaround time, driven not only by pathologist availability but also by upstream coordination and administrative steps (e.g., slide distribution, order entry, and communication across the clinical team). These burdens are amplified in teaching institutions where residents and fellows often contribute substantially to routine preview and ordering. By automating the initial triage step and streamlining IHC ordering, our approach will significantly reduce turnaround time and lower overall costs.

The strong performance of HATS reflects three complementary design choices. First, we leveraged a pretrained pathology foundation model to extract tile-level representations that capture rich, transferable morphologic features. Second, our weakly supervised MIL framework learns how to aggregate these features into slide-level classifications without the need for exhaustive tile-level annotation. This is an especially important advantage in hematopathology, where diffuse infiltration, heterogeneous backgrounds, and subtle patterns make granular labeling both difficult and expensive. Third, the study benefits from scale: this work assembles one of the largest hematopathology tissue WSI cohorts reported for multi-class lymphoma triage, providing both the statistical power and the breadth of morphologic variability needed to train and validate a model intended for real-world clinical deployment.

Importantly, the strong performance of H-optimus-0 in our evaluation suggests that general-purpose pathology foundation models, despite not being trained specifically on hematopathology data, capture sufficient morphological signal for lymphoma triage. This finding has practical significance as it allows institutions to achieve clinically useful performance by combining publicly available foundation models with lightweight downstream classifiers. The rapid iteration this enables was central to our study design, allowing systematic comparison of multiple foundation model architectures and dozens of training configurations at negligible computational cost. This substantially lowers the barrier to entry for any institution seeking to adopt AI-assisted workflows regardless of available computational resources and infrastructure.

In assembling the study cohort, we intentionally prioritized a case mix that reflects real-world hematopathology practice, which necessarily results in an imbalanced class distribution. While we enriched several less common but clinically important entities (e.g., ALCL, AITL, CHL, and BL), we did not upsample them to match the prevalence of the most prevalent categories. This design choice was deliberate: in clinical workflows, “common things are common,” and a triage system must be optimized for the distribution of cases it will encounter day-to-day. Moreover, because the goal of HATS is workflow triaging that improves overall throughput, our emphasis was on maximizing aggregate, population-level performance rather than forcing uniform performance across every individual class at the expense of real-world utility.

Building on the model’s robust subtype discrimination, we translated predictions into a practical IHC ordering strategy aligned with how hematopathology workups are executed in routine care. Specifically, we mapped the ten-class outputs to our institutional triage panels: a high-grade B-cell panel designed to cover DLBCL, FL, and BL, a low-grade B-cell panel to cover CLL, MCL, and MZL, and entity-focused panels for AITL, CHL, ALCL, and PCN. This panel-based triaging improves real-world ordering success because many IHC markers overlap across related differentials; consequently, the “correct” panel often remains appropriate even when the model’s fine-grained subtype prediction is imperfect. Importantly, these gains reflect not only model accuracy but also the workflow and panel design choices of the deploying institution, highlighting that end-to-end performance will depend on local ordering conventions, available markers, and clinical practice patterns, and should be calibrated accordingly during implementation.

In routine clinical practice, triage decisions are not based on morphology alone; they are informed by the clinical history and, when available, preliminary flow cytometry findings and other ancillary studies. Accordingly, an important next step is to extend HATS to incorporate these complementary signals. We plan to add a large language model (LLM)-enabled clinical history review component, as the history often provides strong discriminative information that can materially influence the differential diagnosis and the choice of an initial IHC panel. How best to represent, weight, and audit this information alongside WSI-derived predictions will require careful exploration to ensure interpretability and safety. In parallel, flow cytometry is frequently decisive in hematolymphoid diagnosis; integrating timely preliminary flow results into the triage decision could further improve panel selection and reduce downstream rework. AI-based diagnosis in healthcare faces significant barriers, and even very high accuracy can translate into unacceptable patient risk at scale. For that reason, AI-assisted triage is an ideal starting point for clinical adoption: it delivers immediate operational value without requiring fully autonomous decision-making. In this human-in-the-loop framework, the model processes cases and recommends next-step workups, while pathologists retain full responsibility for the final diagnosis after integrating morphology with IHC, flow cytometry, molecular studies, and clinical history. When the model cannot triage confidently, cases are automatically routed to manual review ensuring safety while still capturing the efficiency benefits of automation. This same triage-first paradigm, “AI to prioritize, humans to decide”, can be readily adapted to many other areas of pathology workflows, and importantly, using readily available pathology foundation models.

## METHODS

### Dataset characteristics

The study cohort comprised 4,996 whole slide images from 1,607 patients representing ten diagnostic categories of hematolymphoid neoplasms: follicular lymphoma (FL, n=1,341 slides), diffuse large B-cell lymphoma (DLBCL, n=1,320), plasma cell neoplasm (PCN, n=537), marginal zone lymphoma (MZL, n=511), mantle cell lymphoma (MCL, n=391), classical Hodgkin lymphoma (CHL, n=324), chronic lymphocytic leukemia/small lymphocytic lymphoma (CLL, n=266), anaplastic large cell lymphoma (ALCL, n=127), angioimmunoblastic T-cell lymphoma (AITL, n=111), and Burkitt lymphoma (BL, n=68). The distribution of diagnostic categories was imbalanced, with FL and DLBCL comprising over 50% of the dataset while AITL and BL represented less than 4% combined. This, however, roughly reflects the distribution of real-world cases. This study was approved by Memorial Sloan Kettering Cancer Center’s Institutional Review Board.

Following patient-level stratification, the dataset was partitioned into training (70%), validation (15%), and test (15%) sets with no patient overlap between partitions. Slides varied substantially in size, with patch counts ranging from 50 to 61,203 (mean 6,343, median 2,164 patches per slide). Slides with fewer than 50 patches were excluded from analysis, and slides exceeding 10,000 patches were randomly subsampled at each training epoch.

### Whole slide image preprocessing

Glass slides were digitized at ×40 equivalent magnification (∼0.25 μm per pixel) on Leica Aperio GT450 (Leica Biosystems, Buffalo Grove, Illinois). Whole slide images (WSIs) were processed using a three-stage computational pipeline implemented with the Trident pathology library[27]. Tissue segmentation was performed using the Grandqc-artifact convolutional neural network[28], which identifies viable tissue regions while excluding background, pen marks, air bubbles, and tissue folds. The segmentation model was applied with a confidence threshold of 0.5, and outputs were stored as GeoJSON contour files for quality control review and downstream processing.

Non-overlapping patches were then extracted from segmented tissue regions at 20× magnification (0.5 μm/pixel) to match foundation model input requirements. A minimum tissue proportion threshold of 0.2 was applied, excluding patches where background content exceeded 80% of the patch area. Patch coordinates were stored in HDF5 format.

Patch-level feature representations were extracted using pathology foundation models: large vision transformers pretrained via self-supervised learning on millions of histopathology images spanning diverse organ systems and disease entities. We evaluated seven foundation models: H-optimus-0 (a 1.1-billion parameter vision transformer trained on over 500,000 WSIs, 1536-dimensional embeddings), Virchow2 (trained on 3.1 million slides, 1024-dimensional), UNI v2 (an updated version of UNI trained on an expanded dataset, 1024-dimensional), Prov-GigaPath (a DINOv2-based vision transformer trained on 1.3 billion image tiles from over 170,000 slides, 1536-dimensional), CONCH (vision-language model trained on 1.17 million image-caption pairs, 512-dimensional), MUSK (multimodal model trained on pathology images and clinical reports, 1024-dimensional), and Phikon v2 (a DINOv2-based model pretrained on TCGA and other public histopathology datasets, 768-dimensional). Each patch was encoded into a fixed-dimensional feature vector capturing morphological patterns, cellular organization, and tissue architecture.

### Multiple-instance learning model

Slide-level classification was performed using a gated attention-based multiple instance learning (MIL) architecture that aggregates patch-level features into slide-level predictions without requiring patch-level annotations. The model treats each slide as a “bag” containing variable numbers of patch “instances” and learns to identify diagnostically relevant regions through a trainable attention mechanism.

The gated attention module computes instance-level importance scores through two parallel transformation pathways. For an input feature vector h_k_ ∈ R^d^ where d=1,536 representing the k-th patch, attention weights are computed as:

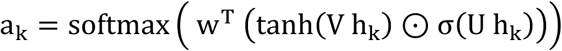

where V, U ∈ R^{c × d}^ (c=128) are learned projection matrices that map input features to a 128-dimensional latent space, w ∈ R^c^ is a learned weight vector that produces scalar attention scores, σ denotes the sigmoid activation function, tanh is the hyperbolic tangent activation, and ⊙ represents element-wise (Hadamard) multiplication. The tanh-activated pathway captures feature relevance for classification, while the sigmoid-activated pathway acts as a learnable gate that modulates information flow by scaling activations between 0 and 1. This gating mechanism enables the model to selectively suppress uninformative features while amplifying discriminative patterns. The slide-level representation *z* ∈ *R*^1024^ is computed as the attention-weighted sum of all patch features:

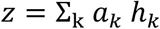

This aggregation produces a fixed-dimensional slide representation regardless of the number of constituent patches. The classification head architecture adapts based on feature dropout rate to provide appropriate model capacity. Although for our final model we chose a constant feature dropout rate, we tested various values. For dropout rates exceeding 0.3, a two-layer architecture was employed: the pooled features pass through a fully-connected layer that projects from 1536 to 512 dimensions, followed by ReLU activation, an additional dropout layer with rate equal to half the primary dropout rate, and a final linear layer projecting to the number of output classes. For lower dropout rates, a single linear layer maps directly from the 1536-dimensional pooled representation to class logits. All weight matrices were initialized using Xavier uniform initialization to promote stable gradient flow during early training epochs, with bias terms initialized to zero.

### Regularization

Multiple complementary regularization strategies were employed to prevent overfitting and improve generalization. Feature dropout with a rate of 0.4 was applied to input patch features prior to attention computation and to the aggregated slide-level representation before classification. This dropout rate encourages the model to learn robust features that do not depend on any single patch or feature dimension. Attention dropout with a rate of 0.1 was also applied to attention scores before softmax normalization, preventing the model from attending exclusively to a small subset of patches and encouraging broader consideration of relevant tissue regions across each slide.

L2 weight regularization (weight decay) with coefficient λ = 0.01 was applied to all model parameters through the optimizer, penalizing large weight magnitudes and encouraging simpler learned representations. The combination of dropout and weight decay provides complementary regularization: dropout acts as an implicit ensemble by training different subnetworks, while weight decay constrains the hypothesis space by penalizing model complexity.

### Data partitioning

The dataset was partitioned at the patient level to prevent data leakage and ensure unbiased performance estimation. Patient medical record numbers (MRNs) served as unique identifiers, ensuring that all slides from the same patient (including multiple tissue sections, levels, or stain variants from the same biopsy) remained within the same partition. The data was divided into training (70%), validation (15%), and test (15%) sets.

An iterative stratified splitting algorithm was employed to identify partitions that maintain balanced class distributions while respecting patient-level boundaries. The algorithm performed up to 100 random patient-level splits and selected the partition that minimized the total deviation from the target class distribution (computed from the overall dataset) across all three subsets. For each candidate split, the algorithm computed the sum of absolute differences between each partition’s class proportions and the overall dataset proportions, selecting the split with minimum total deviation. This approach ensures that rare diagnostic categories are adequately represented in validation and test sets for reliable performance estimation.

### Embedding quality assessment via k-nearest neighbor classification

To evaluate the discriminative quality of foundation model embeddings prior to any supervised training, we performed k-nearest neighbor (k-NN) classification on slide-level representations derived by mean-pooling. For each slide, all patch-level embedding vectors (shape: number of patches × model dimension) were averaged element-wise to produce a single slide-level vector. This uniform aggregation assigns equal weight to all patches and involves no learnable parameters. The resulting training matrix (approximately 3,400 slides × 1,536 dimensions for H-optimus-0) was used to classify held-out validation slides using scikit-learn’s KNeighborsClassifier with cosine distance metric and inverse-distance weighting. We swept k over {1, 3, 5, 10, 20} and selected the value maximizing macro-averaged F1 score. Classification performance was evaluated using accuracy, macro-averaged F1, and macro-averaged ROC AUC computed from distance-weighted class probabilities.

### Instance sampling and batching

Slides with fewer than 50 tissue patches were excluded from analysis to ensure sufficient morphological representation for reliable classification. For slides containing more than 10,000 patches, random subsampling without replacement was performed at each training epoch, selecting exactly 10,000 patches per slide. The random sampling seed was computed deterministically as a function of both the epoch number and slide identifier (via hash function), ensuring that different patch subsets were sampled across epochs while maintaining reproducibility. This epoch-wise resampling strategy effectively augments the training data by exposing the model to diverse patch combinations from large slides throughout training, reducing overfitting to specific patch subsets while maintaining computational tractability.

A bag size stratified batch sampling strategy was implemented to address the substantial variation in slide sizes across the dataset. Slides were assigned to five bins based on patch count quantiles, with bin boundaries determined by the 20th, 40th, 60^th^, and 80th percentiles of the patch count distribution. During training, batches were constructed by sampling slides from within the same size bin, then shuffling the order of batches across bins. This approach groups similarly-sized slides together within each batch, reducing the amount of zero-padding required and improving computational efficiency. Variable-length bags within each batch were padded to the length of the longest bag using zero vectors. The attention mechanism naturally assigns near-zero weights to padded positions due to their uniform feature values, which produce low activation through both the tanh and sigmoid pathways. Training batches were constructed with drop_last=True to prevent single-sample batches that could destabilize batch-dependent operations.

### Model training

The model was trained using the Adam optimizer with β₁ = 0.9, β₂ = 0.999, learning rate of 1×10⁻⁴, and weight decay of 0.01. Cross-entropy loss was used as the objective function.

Training was conducted with a batch size of 32 slides. However, multiple batch sizes were tested with no noticeable performance difference. At each epoch, training data was reshuffled via the stratified batch sampler. After slides were chosen, a max patch parameter sets the maximum number of patches to be used in this training epoch. If a slide has more than the maximum number, large slides are randomly sampled at every epoch to provide varied patch subsets over the course of training. The model checkpoint achieving highest validation accuracy was then saved for final evaluation.

Alternative methods were explored, though none yielded noticeable performance gains, including trying different loss functions, like focal loss, or changing the loss function mid-training. No gain was noted when using variable learning rates, i.e. rates that decrease with epoch (linearly, step-wise, or sinusoidal). Synthetic bag generation was also attempted for rare classes. This involved feature smearing as well as bag resampling from the larger subtype group. After these attempts, we concluded that the main barrier for these rare subtypes is limited sample size, rather than training techniques.

### Case-level aggregation

Clinical diagnostic workflows typically integrate findings from multiple slides per case, including multiple levels or sections from the same biopsy specimen. To reflect this clinical practice, case-level predictions were generated by aggregating slide-level predictions from all slides belonging to the same case, identified by unique case identifiers in the clinical metadata. Four aggregation strategies were implemented and evaluated: (1) Attention-weighted aggregation assigns higher weight to slides exhibiting more confident (focused) attention patterns. For each slide, attention entropy *H_s_* was computed as the negative sum of attention weights multiplied by their logarithms: *H_s_* = −Σ_k_ *a*_k_ log(*a*_k_). Lower entropy indicates more concentrated attention on fewer patches, suggesting higher model confidence. Slide weights were computed as the inverse of entropy plus a stability constant (ε = 0.1), normalized via softmax with temperature parameter τ = 1.0:

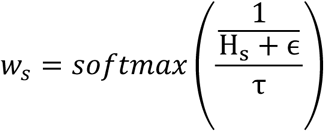

The case-level prediction was computed as the weighted sum of slide-level probability vectors:

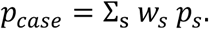

(2) Max pooling aggregation selects the prediction from the slide exhibiting highest classification confidence, measured as the maximum value in the predicted probability vector. This winner-takes-all approach assumes that the most confident slide-level prediction is most likely to reflect the true diagnosis.

(3) Mean pooling aggregation computes the unweighted arithmetic mean of all slide-level probability vectors. This baseline approach treats all slides as equally informative.

(4) Majority voting aggregation assigns each slide a vote for its most probable class (argmax of the probability vector), with the case-level prediction determined by the class receiving the most votes. Ties were resolved using the average probability across tied classes as a tiebreaker.

### Evaluation metrics

Model performance was assessed using a comprehensive suite of metrics computed at both slide-level and case-level granularities.

Classification performance was evaluated using accuracy (proportion of correct predictions), precision (positive predictive value), recall (sensitivity), and F1-score (harmonic mean of precision and recall). For multi-class settings, these metrics were computed per-class and summarized using macro-averaging (unweighted mean across classes, giving equal importance to each class regardless of prevalence) and micro-averaging (pooled across all samples, giving equal importance to each sample).

Discriminative performance was assessed using the area under the receiver operating characteristic curve (ROC AUC), which measures the probability that a randomly chosen positive sample is ranked higher than a randomly chosen negative sample. For multi-class classification, ROC AUC was computed using the one-vs-rest (OVR) strategy, treating each class as positive against all others, with results summarized via macro and micro averaging. The area under the precision-recall curve (AUPRC, also known as average precision) was computed similarly, providing a threshold-independent measure of performance that is particularly informative for imbalanced class distributions where precision at various recall levels is clinically relevant.

Ranking performance was evaluated using top-k accuracy for k ∈ , measuring the proportion of samples where the true class appeared among the k highest-probability predictions. This metric reflects clinical utility in scenarios where multiple differential diagnoses may be considered and the correct diagnosis need not be the single top prediction.

Calibration quality was assessed using expected calibration error (ECE), which measures the alignment between predicted confidence and empirical accuracy. Predictions were grouped into 10 equally-spaced bins by confidence level, and ECE was computed as the weighted average of the absolute difference between mean confidence and mean accuracy within each bin. The Brier score was computed as the mean squared error between predicted probability vectors and one-hot encoded true labels, providing a proper scoring rule that jointly measures calibration and discrimination.

The Matthews correlation coefficient (MCC) was computed as a balanced measure of classification quality that accounts for all four confusion matrix categories and remains informative even with imbalanced class distributions. Confusion matrices were generated to visualize prediction patterns across all class pairs, with row-wise normalization showing the distribution of predictions for each true class.

For case-level evaluation, all metrics were computed using case-level predictions and labels. Additionally, slide-level top-k statistics were computed to assess the proportion of slides per case where the correct diagnosis appeared among the top-k predictions, providing insight into the consistency of slide-level predictions within each case.

## Author contributions

IS and MH developed the computational pipeline, trained the models, and performed the analyses. PG provided expert hematopathology review and defined the IHC panel ordering rules. CV, AK, GG, HV, and JJ provided guidance on model architecture and computational methodology. OA and LG assisted with whole-slide image processing and infrastructure. AL, MZ and AD jointly conceived and supervised the study. IS and MZ wrote the manuscript. All authors reviewed and approved the final manuscript.

## Data availability

HATS model checkpoints are publicly available via HuggingFace at https://huggingface.co/isafa/HATS. The clinical datasets and digital whole-slide images used in this study were generated at Memorial Sloan Kettering Cancer Center and are not publicly available due to institutional policies governing patient privacy.

## Code availability

Code used in this work is available via GitHub at https://github.com/ibsafa/MILTraining

## Supplementary Information

### Supplementary Note 1: Detailed Foundation Model Comparison for Hematologic Malignancy Classification

We evaluated seven publicly available pathology foundation models using identical downstream classifiers and multi-seed experiments to provide a systematic comparison for the hematopathology community. All models were evaluated using frozen encoders (no fine-tuning of foundation model weights), meaning that downstream performance reflects the quality of morphological representations learned during pretraining. Below we describe the models, their characteristics, and detailed performance analysis.

Model characteristics. The seven models span a range of architectures, training scales, and intended applications. H-optimus-0 (Bioptimus; ViT-g, 1.1 billion parameters, 1536-dimensional embeddings, Apache 2.0 license) is the largest model in our comparison, pretrained on over 500,000 whole-slide images from 4,000 clinical practices. Virchow2 (Paige AI; ViT-H, 632 million parameters, 1024-dimensional, CC-BY-NC-ND-4.0) was trained on the largest dataset: 3.1 million WSIs from over 800 laboratories spanning nearly 200 tissue types. UNI v2 (Harvard/MGH; ViT-L, approximately 307 million parameters, 1024-dimensional, CC-BY-NC-ND-4.0) was trained on an expanded dataset of institutional slides. Prov-GigaPath (Microsoft; DINOv2-based with LongNet, 1536-dimensional, Apache 2.0) was pretrained on 1.3 billion image tiles from 171,000 WSIs in the Providence healthcare system. CONCH (Harvard; CoCa vision-language architecture, approximately 90 million vision parameters, 512-dimensional, CC-BY-NC-ND-4.0) was jointly trained on 1.17 million pathology image-caption pairs. MUSK (Stanford; BEiT3-based multimodal, 1024-dimensional, CC-BY-NC-ND-4.0) aligns pathology images with clinical text for precision oncology applications. Phikon v2 (Owkin; ViT-L DINOv2, 768-dimensional, non-commercial license) is the only model in our comparison trained exclusively on publicly available data from TCGA, CPTAC, and GTEx.

Overall performance. H-optimus-0 achieved the best overall performance with a mean slide-level accuracy of 79.4% +/- 1.5% (SD, n=10), case-level accuracy of 83.8% +/- 1.5%, and ROC AUC of 0.962 +/- 0.009. Prov-GigaPath achieved slide-level accuracy of 75.3% +/- 2.8% (SD, n=10), case-level accuracy of 78.3% +/- 2.9%, and ROC AUC of 0.949 +/- 0.015. UNI v2 achieved slide-level accuracy of 77.0% +/- 2.6% (SD, n=10), case-level accuracy of 79.4% +/- 2.3%, and ROC AUC of 0.955 +/- 0.012. Virchow2 achieved slide-level accuracy of 76.7%+/- 2.8% (SD, n=16), case-level accuracy of 80.0% +/- 2.9%, and ROC AUC of 0.955 +/- 0.010. CONCH achieved slide-level accuracy of 71.0% +/- 3.1% (SD, n=18), case-level accuracy of 73.9% +/-3.3%, and ROC AUC of 0.942 +/- 0.009. MUSK achieved slide-level accuracy of 69.3% +/-2.2% (SD, n=10), case-level accuracy of 72.7% +/- 3.4%, and ROC AUC of 0.929 +/- 0.009. Phikon v2 achieved slide-level accuracy of 68.2% +/- 2.5% (SD, n=10), case-level accuracy of 71.1% +/- 2.5%, and ROC AUC of 0.928 +/- 0.011.

Per-class performance. Performance varied substantially by diagnostic category, with common subtypes classified reliably across all models and rare subtypes showing the largest model-dependent variation. At the case level (attention-weighted aggregation), FL accuracy ranged from 83.6% (MUSK) to 91.2% (H-optimus-0), DLBCL from 82.2% (MUSK) to 89.8% (H-optimus-0), and PCN from 84.5% (Phikon v2) to 94.8% (H-optimus-0). Among rare entities, AITL ranged from 0% (Phikon v2) to 49.8% (H-optimus-0), ALCL from 22.1% (Phikon v2) to 50.5% (H-optimus-0), and BL from 0% (Phikon v2) to 48.2% (H-optimus-0). CLL showed a particularly wide range from 41.2% (Phikon v2) to 84.1% (H-optimus-0), and MCL from 32.8% (Phikon v2) to 66.2% (H-optimus-0) (Supplementary Table 1; Supplementary Fig. 2).

Vision-only versus vision-language and multimodal models. The five purely vision-based self-supervised models achieved a mean slide-level accuracy of 75.3%, compared to 71.0% for CONCH (vision-language) and 69.3% for MUSK (multimodal). Because our evaluation pipeline uses only the visual encoder with frozen weights, the text-aligned components of these models are not utilized during feature extraction or classification. The lower performance of CONCH and MUSK in our setting therefore reflects the expected behavior of evaluating only one modality of a multi-modal model, rather than a limitation of these architectures.

Pretraining data and transferability. None of the seven models were specifically designed or pretrained for hematologic malignancy classification, and the public datasets underlying several models (most notably TCGA, with only 48 DLBCL cases among 33 cancer types) contain minimal lymphoid tissue. Importantly, even the models with the least exposure to hematologic tissue achieved accuracy far exceeding chance, demonstrating that general-purpose self-supervised pretraining captures broadly transferable morphological features.

Model consistency. H-optimus-0 demonstrated the lowest performance variance across training seeds (SD = 1.5 percentage points, range 77.2% to 81.3%), followed by MUSK (2.2 pp), Phikon v2 (2.5 pp), UNI v2 (2.6 pp), Prov-GigaPath (2.8 pp), Virchow2 (2.8 pp), and CONCH (3.1 pp). Lower variance indicates that the model’s performance generalizes more reliably across different data partitions and random initializations, with less sensitivity to the specific training conditions.

Taken together, these results support the selection of H-optimus-0 as the default backbone for HATS based on its highest overall performance, lowest variance across seeds, and strong per-class results across both common and rare subtypes.

### Supplementary Note 2: Embedding Quality Assessment via k-Nearest Neighbor Classification

To assess the intrinsic performance of the foundation model embeddings independent of any learned classifier, we performed a parameter-free k-nearest neighbor (k-NN) analysis using H-optimus-0 representations. Each slide’s patch-level embeddings were mean-pooled into a single fixed-dimensional vector (approximately 3,400 slides × 1,536 dimensions), producing a compact slide-level representation without any learned aggregation. Using cosine distance and inverse-distance weighted voting with scikit-learn’s KNeighborsClassifier, we swept k over {1, 3, 5, 10, 20} and selected the value maximizing macro-averaged F1 score.

The k-NN classifier achieved 68% slide-level accuracy on the held-out validation set, well above the 10% expected by chance for a ten-class problem. This result demonstrates that the frozen H-optimus-0 embeddings place same-subtype slides near each other in the representation space even before any task-specific training, confirming that the foundation model captures intrinsic morphological distinctions among lymphoma subtypes. The subsequent addition of the gated attention mechanism through ABMIL training then pushes overall accuracy from 68% to 79% at the slide level.

Per-class analysis (Supplementary Fig. 3) reveals that the ABMIL improvement is not uniform across subtypes. Common subtypes with abundant training data (PCN, FL, DLBCL) show relatively modest gains from k-NN to ABMIL, as the k-NN baseline is already strong for these classes. In contrast, rare subtypes (BL, ALCL, AITL) show the largest absolute improvements, indicating that the attention mechanism is particularly important for learning discriminative patterns from limited training examples. However, these rare classes also exhibit the largest variance across training seeds, consistent with the limited statistical power available for these categories. The improvement from k-NN to ABMIL for CHL is notably large, suggesting that CHL morphology may be better captured by learned attention patterns that focus on diagnostic features (such as Reed-Sternberg cells) rather than by global mean-pooled representations.

### Supplementary Note 3: Performance Stratified by Tissue Type

To assess whether HATS performance depends on the anatomical source of the tissue specimen, we stratified model predictions by tissue type. Slides were classified as lymph node or non-lymph node specimens based on the tissue part description extracted from institutional pathology reports. This yielded 400 lymph node slides and 266 non-lymph node slides per seed (across 10 independent training seeds with different data partitions).

Lymph node specimens achieved a mean slide-level accuracy of 78.7% ± 2.9% and macro-averaged F1 score of 0.658 ± 0.067 across seeds. Non-lymph node specimens achieved a mean accuracy of 79.9% ± 3.9% and macro-averaged F1 score of 0.669 ± 0.090 (Supplementary Fig. 4a,b). The difference in accuracy between tissue types was not statistically significant (paired t-test, t = −0.675, p = 0.516; Mann–Whitney U = 41.0, p = 0.521), indicating that model performance is comparable across lymph node and non-lymph node specimens.

Per-class analysis revealed tissue-type-dependent variation that reflects the underlying biology of the disease subtypes (Supplementary Fig. 4c). The most striking difference was observed for MZL, where accuracy in non-lymph node specimens (83.6% ± 8.5%) was markedly higher than in lymph node specimens (40.4% ± 20.4%), consistent with the predominantly extranodal nature of MZL, the majority of which arises in mucosa-associated lymphoid tissue (MALT) sites.

Among the remaining subtypes, a similar trend was apparent, though wide standard deviations across training seeds limit the strength of these comparisons. Subtypes that predominantly arise in lymph nodes, such as FL (lymph node: 89.6% ± 3.2%, non-lymph node: 84.8% ± 5.0%) and CHL (80.1% ± 5.0% vs. 68.8% ± 29.5%), tended toward higher accuracy in lymph node specimens, whereas subtypes that frequently present in extranodal sites, including DLBCL (79.8% ± 7.8% vs. 88.7% ± 5.0%) and PCN (93.1% ± 11.4% vs. 85.8% ± 9.6%), showed a modest advantage in non-lymph node tissue. MCL performance was similarly higher in lymph node specimens (74.0% ± 12.0% vs. 60.5% ± 13.4%).

Prediction confidence was comparable between tissue types, with both lymph node and non-lymph node specimens achieving a mean confidence of 0.842 (Mann–Whitney U, p = 0.146; Supplementary Fig. 4d). Unlike the previous Virchow2-based analysis, where non-lymph node specimens showed slightly higher confidence, H-optimus-0 shows no significant difference in prediction confidence across tissue sources.

Together, these results demonstrate that HATS achieves consistent performance across anatomical tissue sources. The absence of a significant accuracy difference between lymph node and non-lymph node specimens supports the generalizability of the model across the diversity of specimen types encountered in routine hematopathology practice.

**Supplementary Table 1.** Foundation model comparison. Slide-level and case-level classification performance across seven pathology foundation models. Each model was evaluated with 10 to 20 independent training seeds using identical MIL classifier architecture and hyperparameters. Values represent mean ± standard deviation.

| Model | Slide Accuracy | F1 (Macro) | Case Accuracy |
| --- | --- | --- | --- |
| **H-optimus-0** | $0.794 \pm 0.015$ | $0.708 \pm 0.037$ | $0.838 \pm 0.015$ |
| **Virchow2** | $0.767 \pm 0.028$ | $0.661 \pm 0.059$ | $0.800 \pm 0.029$ |
| UNI V2 | $0.770 \pm 0.026$ | $0.670 \pm 0.062$ | $0.794 \pm 0.023$ |
| Prov-GigaPath | $0.753 \pm 0.028$ | $0.641 \pm 0.062$ | $0.783 \pm 0.029$ |
| CONCH | $0.710 \pm 0.031$ | $0.555 \pm 0.058$ | $0.739 \pm 0.033$ |
| MUSK | $0.693 \pm 0.022$ | $0.550 \pm 0.048$ | $0.727 \pm 0.034$ |
| Phikon v2 | $0.682 \pm 0.025$ | $0.466 \pm 0.039$ | $0.711 \pm 0.025$ |

*†Paired t-test comparing per-seed slide-level accuracy distributions against H-optimus-0 (reference). \* $p < 0.05$ ; \*\*\* $p < 0.001$ . Case accuracy computed using attention-weighted aggregation.*

**Supplementary Table 2.** Per-class case-level accuracy across foundation models. Mean case-level accuracy (attention-weighted aggregation) per lymphoma subtype across independent training seeds for each foundation model. Bold values indicate the highest accuracy for each class.

| Model | AITL | ALCL | BL | CHL | CLL | DLBCL | FL | MCL | MZL | PCN |
| --- | --- | --- | --- | --- | --- | --- | --- | --- | --- | --- |
| H-optimus-0 | 0.498 | 0.505 | 0.482 | 0.802 | 0.841 | 0.898 | 0.912 | 0.662 | 0.741 | 0.948 |
| Virchow2 | 0.323 | 0.434 | 0.419 | 0.766 | **0.775** | 0.885 | **0.904** | 0.584 | **0.687** | **0.902** |
| UNI V2 | 0.339 | **0.443** | **0.428** | **0.778** | 0.630 | **0.886** | 0.901 | **0.641** | 0.651 | 0.897 |
| Prov-GigaPath | 0.334 | 0.475 | 0.438 | 0.729 | 0.698 | 0.820 | 0.826 | 0.638 | 0.688 | 0.891 |
| CONCH | 0.205 | 0.227 | 0.235 | 0.802 | 0.644 | 0.850 | 0.862 | 0.511 | 0.469 | 0.907 |
| MUSK | 0.082 | 0.335 | 0.400 | 0.773 | 0.638 | 0.822 | 0.771 | 0.578 | 0.540 | 0.806 |
| Phikon v2 | 0.000 | 0.221 | 0.000 | 0.691 | 0.412 | 0.892 | 0.879 | 0.328 | 0.521 | 0.845 |

Classes ordered alphabetically. AITL, angioimmunoblastic T-cell lymphoma; ALCL, anaplastic large cell lymphoma; BL, Burkitt lymphoma; CHL, classical Hodgkin lymphoma; CLL, chronic lymphocytic leukemia; DLBCL, diffuse large B-cell lymphoma; FL, follicular lymphoma; MCL, mantle cell lymphoma; MZL, marginal zone lymphoma; PCN, plasma cell neoplasm.

**Supplementary Table 3.** IHC panel composition. Immunohistochemical stains included in each diagnostic panel recommended by HATS. Panel assignment is determined by the model’s top predicted lymphoma subtype.

| Name | Immunohistochemical Stains |
| --- | --- |
| High-Grade B-Cell Panel (HGBCL) | CD20, CD19, CD3, CD10, BCL2, BCL6, MUM1, MYC, Ki67, CD23, CD21 |
| Low-Grade B-Cell Panel (LGBCL) | CD20, CD3, CD10, BCL2, BCL6, MUM1, Ki67, CD23, CD5, CD138, Kappa, Lambda, LEF1, Cyclin D1 |
| Subtype-Specific (CHL) | CD15, CD30, CD20, CD3, CD45, Pax5, LMP1, MUM1 |
| Subtype-Specific (AITL) | CD2, CD3, CD4, CD5, CD7, CD8, CD10, CD20, CD21, CD23, CD30, CXCL13, PD1, ICOS, EBER, TCR $\alpha$, TCR $\beta$, TRBC1, Ki67 |
| Subtype-Specific (ALCL) | CD2, CD3, CD4, CD5, CD7, CD8, CD20, TCR $\alpha$, TCR $\beta$, TCR $\delta$, CD56, CD30, ALK, TIA1, GzB, P63 |
| Subtype-Specific (PCN) | CD138, BCMA, Kappa-ISH, Lambda-ISH, CD56, CD117, CyclinD1 |

**Supplementary Table 4.** Performance by tissue type. Slide-level classification performance stratified by tissue type across 10 independent training seeds. Values represent mean ± standard deviation.

| Tissue Type | N slides | Accuracy | F1 (Macro) |
| --- | --- | --- | --- |
| Lymph node | 400 | 0.787 ± 0.029 | 0.658 ± 0.067 |
| Non-lymph node | 266 | 0.799 ± 0.039 | 0.669 ± 0.090 |

**Supplementary Figure 1.**
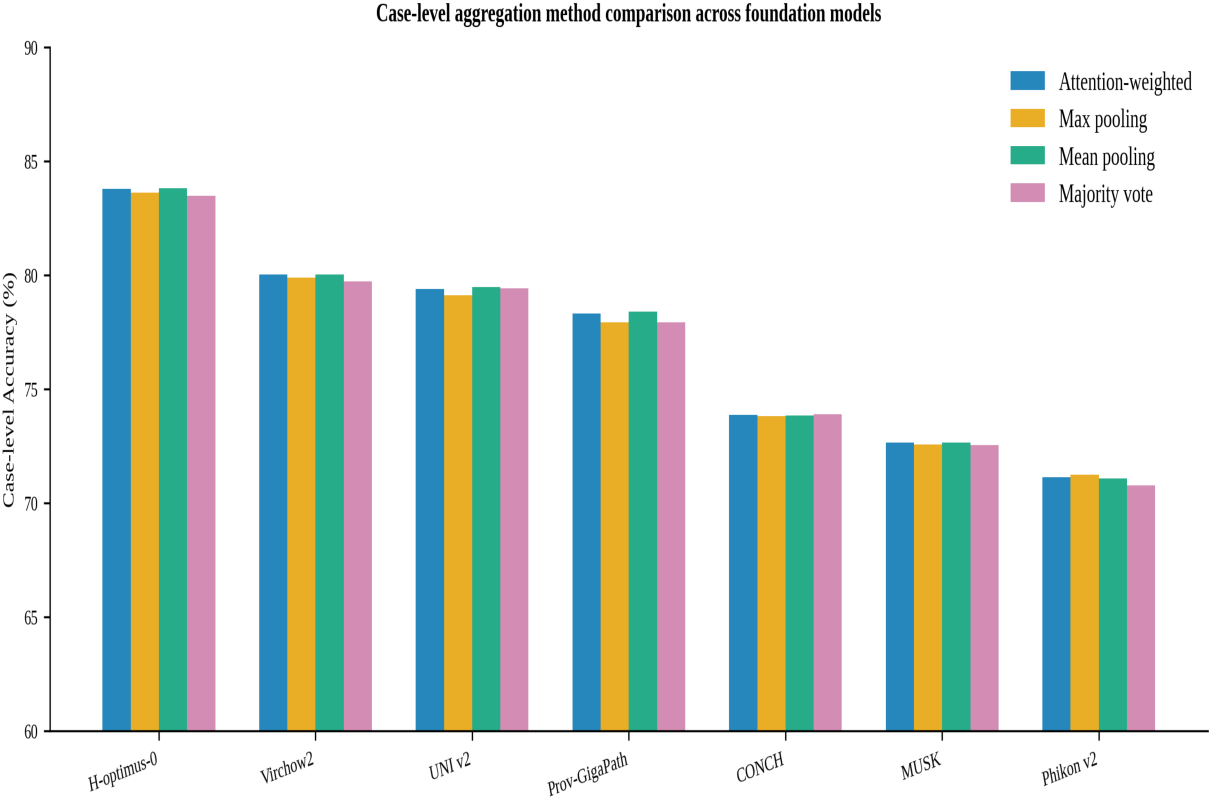
Case-level aggregation method comparison across foundation models. Bar chart showing case-level accuracy (%) for each of seven foundation models (H-optimus-0, Virchow2, UNI v2, Prov-GigaPath, CONCH, MUSK, and Phikon v2) under four aggregation strategies: attention-weighted, max pooling, mean pooling, and majority voting. All methods yielded comparable performance, with minimal variation across aggregation strategies within each model.

**Supplementary Figure 2.**
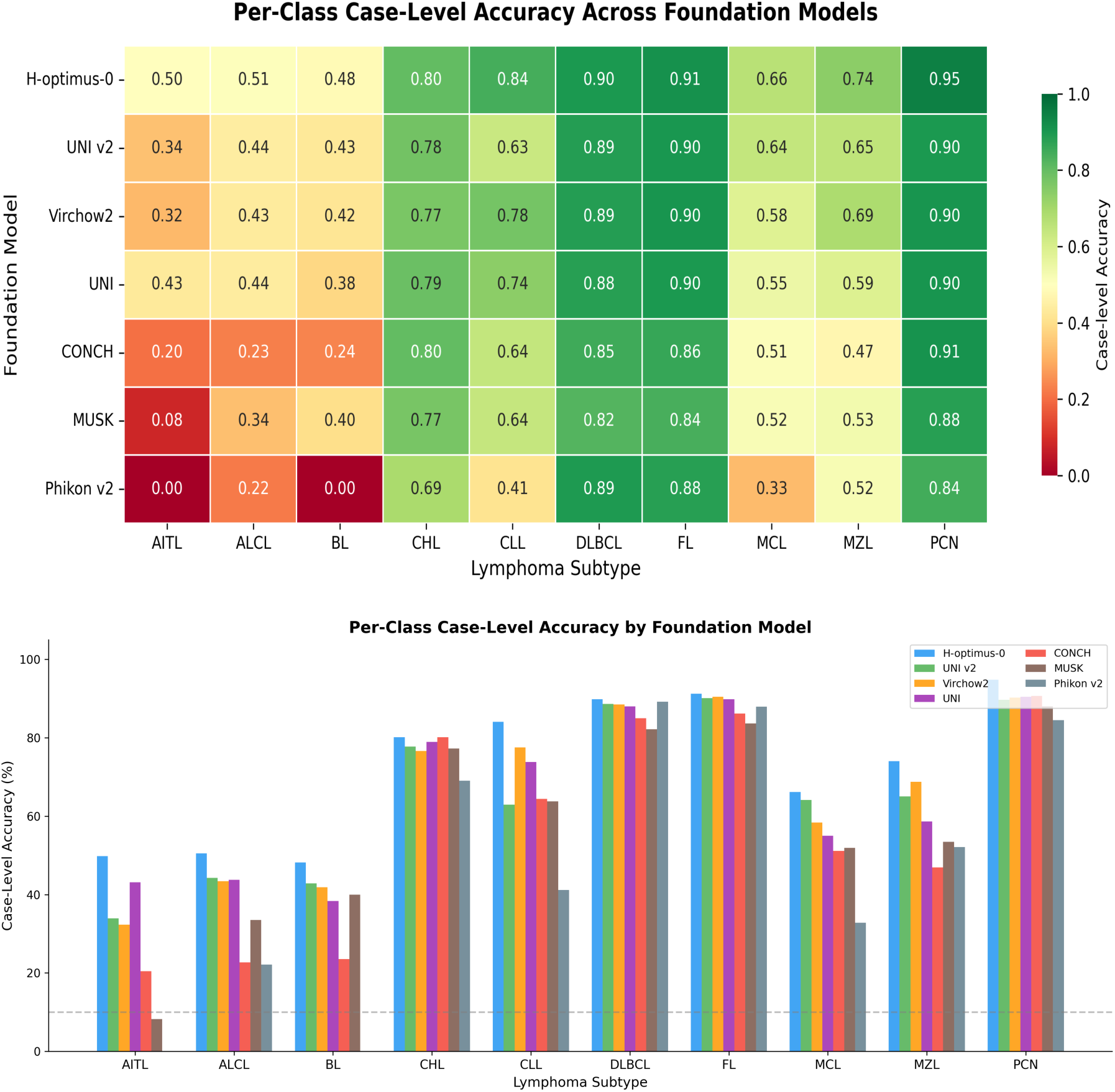
Per-class performance comparison across foundation models. **(a)** Heatmap of per-class F1 scores for each foundation model. Color intensity indicates F1 score magnitude. **(b)** Heatmap of per-class accuracy (recall) for each foundation model. Higher values (green) indicate better classification for that subtype. Both panels demonstrate that common subtypes (FL, PCN, DLBCL) achieve consistently high performance across models, while rare subtypes (AITL, ALCL, BL) show substantial inter-model variability.

**Supplementary Figure 3.**
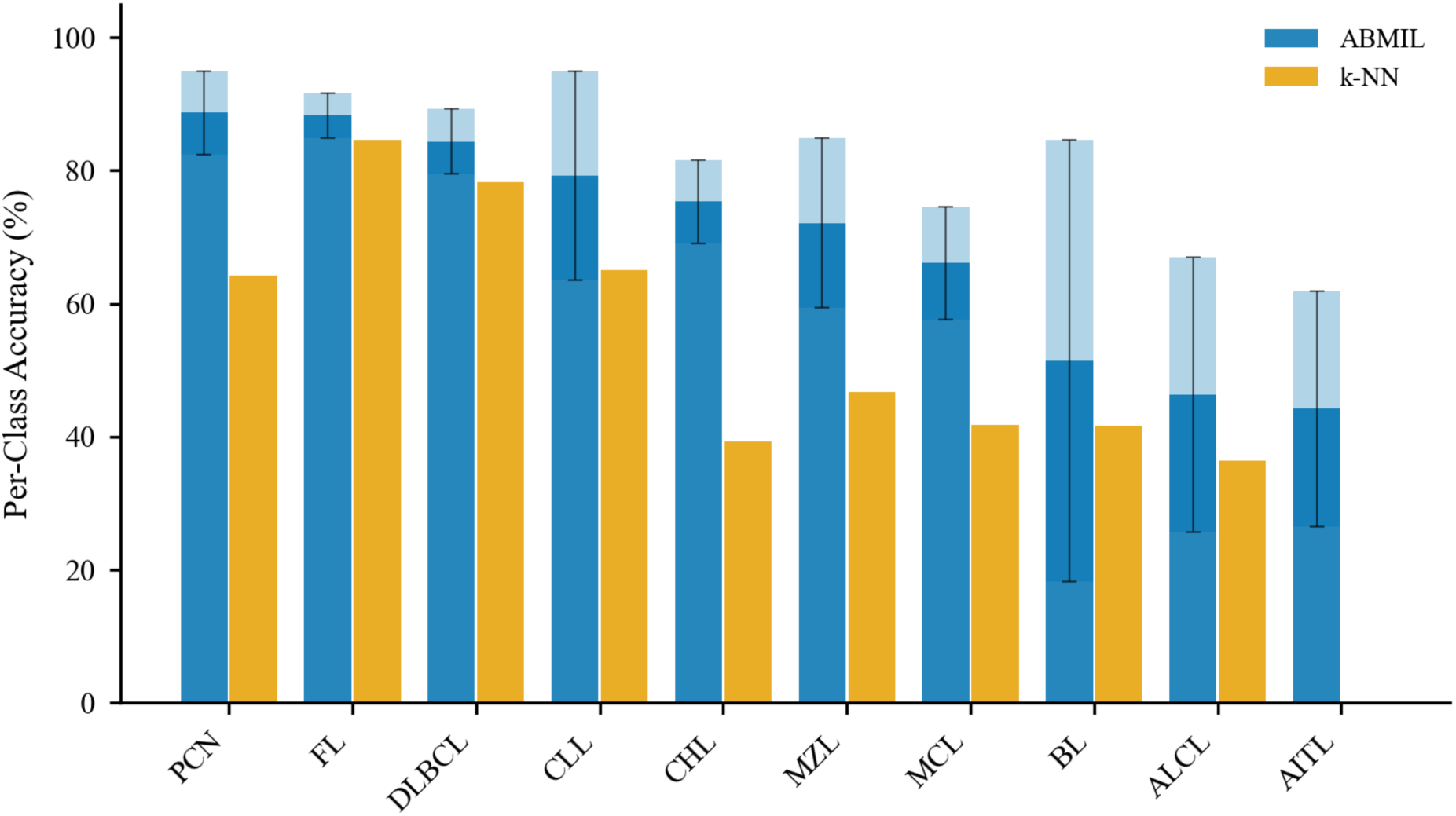
Per-class accuracy comparison between k-NN baseline and ABMIL classifier using H-optimus-0 embeddings. Gold bars indicate zero-shot k-NN accuracy using mean-pooled slide embeddings with cosine distance. Blue bars indicate ABMIL classifier accuracy after training. Light blue extensions show one standard deviation across independent training seeds. Classes are ordered by prevalence in the dataset (left to right: most common to rarest). The ABMIL classifier improves accuracy across all subtypes, with the largest gains observed for rare entities and CHL, where learned attention patterns provide substantial benefit over uniform mean-pooling.

**Supplementary Figure 4.**
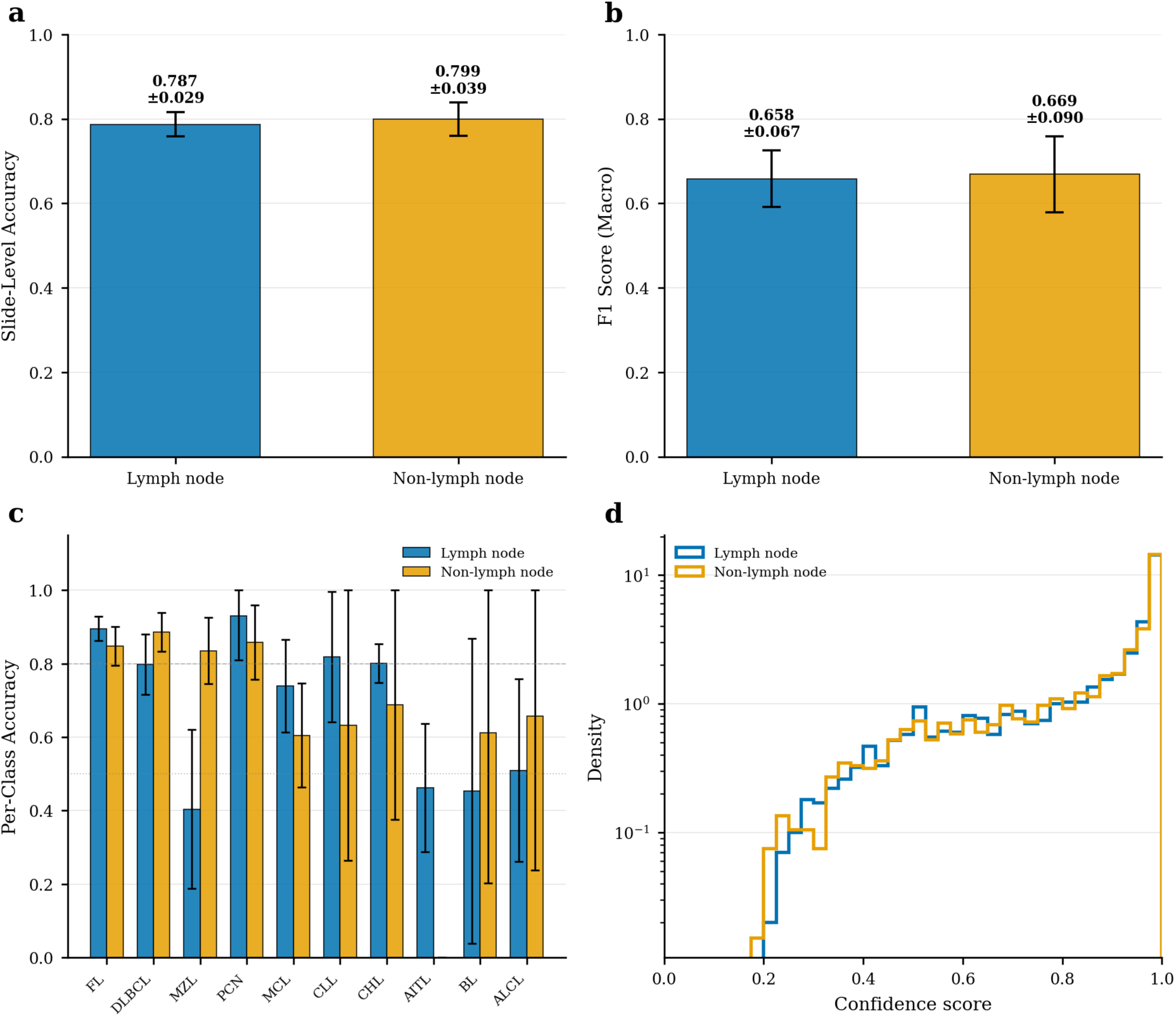
HATS performance stratified by tissue type. **(a)** Slide-level accuracy for lymph node (n = 400) and non-lymph node (n = 266) specimens. Bars indicate mean accuracy across 10 independent training seeds; error bars indicate standard deviation.**(b)** Macro-averaged F1 score by tissue type. **(c)** Per-class accuracy comparison between lymph node and non-lymph node specimens. Classes are ordered by prevalence in the test set. Error bars indicate standard deviation across seeds. **(d)** Distribution of prediction confidence scores by tissue type, shown as step histograms on a logarithmic scale. The overlapping distributions indicate comparable model confidence across lymph node and non-lymph node specimens.

